# RISK FACTORS FOR INFECTION AND HEALTH IMPACTS OF THE COVID-19 PANDEMIC IN PEOPLE WITH AUTOIMMUNE DISEASES

**DOI:** 10.1101/2021.02.03.21251069

**Authors:** Kathryn C. Fitzgerald, Christopher A. Mecoli, Morgan Douglas, Samantha Harris, Berna Aravidis, Jemima Albayda, Elias S. Sotirchos, Ahmet Hoke, Ana-Maria Orbai, Michelle Petri, Lisa Christopher-Stine, Alan N. Baer, Julie J. Paik, Brittany L. Adler, Eleni Tiniakou, Homa Timlin, Pavan Bhargava, Scott D. Newsome, Arun Venkatesan, Vinay Chaudhry, Thomas E. Lloyd, Carlos A. Pardo, Barney J. Stern, Mark Lazarev, Brindusa Truta, Shiv Saidha, Edward S. Chen, Michelle Sharp, Nisha Gilotra, Edward K. Kasper, Allan C. Gelber, Clifton O. Bingham, Ami A. Shah, Ellen M. Mowry

**Affiliations:** Department of Neurology, Johns Hopkins School of Medicine, Baltimore, MD, USA; Department of Epidemiology, Johns Hopkins Bloomberg School of Public Health, Baltimore, MD, USA; Division of Rheumatology, Department of Medicine, Johns Hopkins School of Medicine, Baltimore, MD, USA; Division of Gastroenterology and Hepatology, Department of Medicine, Johns Hopkins School of Medicine, Baltimore, MD, USA; Division of Pulmonary and Critical Care Medicine, Johns Hopkins School of Medicine, Baltimore, MD, USA; Department of Cardiology, Johns Hopkins School of Medicine, Baltimore, MD, USA

## Abstract

**Background:** People with autoimmune or inflammatory conditions who take immunomodulatory/suppressive medications may have a higher risk of novel coronavirus disease 2019 (COVID-19). Chronic disease care has also changed for many patients, with uncertain downstream consequences.

**Objective:** Assess whether COVID-19 risk is higher among those on immunomodulating or suppressive agents and characterize pandemic-associated changes to care.

**Design:** Longitudinal registry study

**Participants:** 4666 individuals with autoimmune or inflammatory conditions followed by specialists in neurology, rheumatology, cardiology, pulmonology or gastroenterology at Johns Hopkins

**Measurements:** Periodic surveys querying comorbidities, disease-modifying medications, exposures, COVID-19 testing and outcomes, social behaviors, and disruptions to healthcare

**Results:** A total of 265 (5.6%) developed COVID-19 over 9 months of follow-up (April-December 2020). Patient characteristics (age, race, comorbidity, medication exposure) were associated with differences in social distancing behaviors during the pandemic. Glucocorticoid exposure was associated with higher odds of COVID-19 in multivariable models incorporating behavior and other potential confounders (OR: 1.43; 95%CI: 1.08, 1.89). Other medication classes were not associated with COVID-19 risk. Diabetes (OR: 1.72; 95%CI: 1.08, 2.73), cardiovascular disease (OR: 1.68; 95%CI: 1.24, 2.28), and chronic kidney disease (OR: 1.76; 95%CI: 1.04, 2.97) were each associated with higher odds of COVID-19. Pandemic-related disruption to care was common. Of the 2156 reporting pre-pandemic utilization of infusion, mental health or rehabilitative services, 975 (45.2%) reported disruptions. Individuals experiencing changes to employment or income were at highest odds of care disruption.

**Limitations:** Results may not be generalizable to all patients with autoimmune or inflammatory conditions. Information was self-reported.

**Conclusions:** Exposure to glucocorticoids may increase risk of COVID-19 in people with autoimmune or inflammatory conditions. Disruption to healthcare and related services was common. Those with pandemic-related reduced income may be most vulnerable to care disruptions.

## INTRODUCTION

In people with autoimmune or inflammatory conditions, there is concern that immunomodulatory medications used to treat these conditions may increase the risk of developing novel coronavirus disease 2019 (COVID-19), as chronic immune suppression is associated with a higher risk of infection and may be associated with poorer COVID-19 outcomes.^1–7^ Given the significant role of immune cell activation and inflammation in severe COVID-19 disease, certain classes of these medications may also have a protective role in this setting. A notable limitation of many existing studies is a failure to account for social distancing behaviors (e.g., individuals taking stronger immune-modulating/suppressive medications may perceive themselves to be at a higher COVID-19 risk and may be more likely to strictly adhere to suggested social distancing guidelines and refrain from in-person socialization). Such differences in behavior could potentially confound (or impact in unexpected ways) any estimates of medication-associated risks. As a result, consideration of behavior remains a critical component in analyses assessing COVID-19 risk associated with exposure to immunomodulatory/suppressive medications.

Beyond COVID-19 disease-specific concerns, the pandemic has resulted in the disruption of longitudinal care in many chronic conditions. For example, many patients have discontinued, lowered, or delayed their medication or refrained from obtaining critical safety laboratory tests.^8,9^ Other patients experienced disruptions to infusion, rehabilitative, homecare, or mental health services. Currently, limited research has assessed 1) which patients are most vulnerable to care disruption and 2) the downstream effects of these disruptions on disease outcomes. The impact of such changes may be very significant as they may affect the majority of patients with autoimmune or inflammatory conditions, not only those patients who develop COVID-19.

To address some of these critical gaps in knowledge and patient care, we established COVID-19 Risk with Immune-modulating Medication Study (COVID-RIMS), which is a cohort study of nearly 5000 individuals with a variety of autoimmune and inflammatory conditions who are managed as outpatients by specialists at the Johns Hopkins Medical Institutions. The primary goals of COVID-RIMS were to assess whether the risk for COVID-19 is higher among those on immunomodulating or suppressive agents (after accounting for behavior) and to quantify and identify consequences of pandemic-associated changes to longitudinal outpatient care in this population.

## METHODS

### Study population

In April 2020, we established the COVID-RIMS research study. Eligible participants have autoimmune or inflammatory conditions (included diagnoses listed in **eTable 1)** and had been seen by specialists in neurology, rheumatology, cardiology, pulmonology or gastroenterology at Johns Hopkins since 2017. Participants were identified as those with at least two ICD-10 codes associated with a particular disorder in the electronic medical record (EMR) at two separate visits prior to study initiation. Patients were invited to participate in COVID-RIMS through a patient-specific e-mail link. This study was conducted following the International Conference on Harmonization guidelines for Good Clinical Practices, the Declaration of Helsinki, as well as local requirements. Approval was obtained from the Institutional Review Board at Johns Hopkins Medical Institutions.

**Table 1.** Characteristics of COVID-RIMS Participant by COVID-19 status.

|  | Overall | No COVID-19 | COVID-19+ | P value* |
| --- | --- | --- | --- | --- |
| N | 4666 | 4401 | 265 |  |
| Age | 55.10 (13.77) | 55.28 (13.77) | 52.19 (13.46) | <0.001 |
| Male sex | 1086 (23.3) | 1036 (23.5) | 50 (18.9) | 0.094 |
| Smoker | 167 (3.6) | 159 (3.6) | 8 (3.0) | 0.737 |
| **Race** |  |  |  | 0.212 |
| White | 3877 (83.1) | 3662 (83.2) | 215 (81.1) |  |
| Asian | 125 (2.7) | 122 (2.8) | 3 (1.1) |  |
| Black or African American | 447 (9.6) | 413 (9.4) | 34 (12.8) |  |
| Other | 171 (3.7) | 161 (3.7) | 10 (3.8) |  |
| Unknown | 45 (1.0) | 42 (1.0) | 3 (1.1) |  |
| Hispanic or Latino | 146 (3.1) | 136 (3.1) | 10 (3.8) | 0.661 |
| In person socializing over follow-up | 2737 (58.7) | 2559 (58.1) | 178 (67.2) | 0.005 |
| In person socializing at baseline | 851 (18.2) | 798 (18.1) | 53 (20.0) | 0.495 |
| Employment status change due to COVID-19 pandemic | 548 (11.7) | 498 (11.3) | 50 (18.9) | <0.001 |
| Working onsite | 1138 (24.4) | 1041 (23.7) | 97 (36.6) | <0.001 |
| Change in ability to pay for care associated costs | 608 (13.0) | 546 (12.4) | 62 (23.4) | <0.001 |
| BMI | 29.51 (7.45) | 29.47 (7.39) | 30.23 (8.33) | 0.116 |
| **Comorbidity** |  |  |  |  |
| Stroke | 159 (3.4) | 150 (3.4) | 9 (3.4) | 1.000 |
| Asthma | 748 (16.0) | 696 (15.8) | 52 (19.6) | 0.120 |
| Cardiovascular disease (CVD) | 877 (18.8) | 814 (18.5) | 63 (23.8) | 0.040 |
| Hypertension | 1437 (30.8) | 1362 (30.9) | 75 (28.3) | 0.402 |
| Chronic kidney disease (CKD) | 195 (4.2) | 178 (4.0) | 17 (6.4) | 0.086 |
| Diabetes | 283 (6.1) | 261 (5.9) | 22 (8.3) | 0.150 |
| Cancer | 544 (11.7) | 515 (11.7) | 29 (10.9) | 0.783 |
| Lung | 520 (11.1) | 485 (11.0) | 35 (13.2) | 0.318 |
| Number of comorbidities | 1.02 (1.14) | 1.01 (1.13) | 1.14 (1.22) | 0.079 |
| Number of autoimmune conditions | 1.33 (0.66) | 1.33 (0.65) | 1.40 (0.83) | 0.083 |
| Ever treated with immune modulating medication | 4161 (89.2) | 3915 (89.0) | 246 (92.8) | 0.062 |
| **Changes to care** |  |  |  |  |
| Any disruption in care/services | 975 (45.2) | 903 (44.6) | 72 (55.0) | 0.026 |
| Delay in infusion | 341 (29.4) | 315 (28.7) | 26 (41.9) | 0.038 |
| Delay in rehab services | 623 (57.6) | 575 (57.2) | 48 (63.2) | 0.373 |
| Delay in mental health services | 211 (28.9) | 189 (27.9) | 22 (40.7) | 0.065 |
| Delay in home care services | 65 (25.6) | 58 (24.7) | 7 (36.8) | 0.371 |

\*P values are derived from univariate generalized linear models using a univariate test for differences between COVID-19 cases versus those with no reported evidence of COVID-19.

### Survey assessments

From April through June 2020, participants in COVID-RIMS answered weekly online questionnaires, and from July 2020 onwards, participants completed surveys on an approximately monthly basis. Surveys queried immunomodulatory medication exposure (both current and previous), COVID-19-related symptoms, testing status and outcomes, weight and height (used to calculate body mass index [BMI] as kg/m^2^), comorbid conditions, smoking status, employment (status and location: onsite versus remote), personal and household contact social distancing practices, mask use, and socio-economic status (SES) indicators. Indicators of SES included the area deprivation index (ADI), which is an established composite index incorporating 17 measures of SES derived using geo-coded addresses; nationwide indices range from 0 (least disadvantaged) to 100 (most disadvantaged).^10,11^ Surveys also assessed whether participants had cancelled or postponed infusion visits, rehabilitative, homecare or mental health services and the reason for the change (lack of available appointments, loss of insurance, COVID-19 exposure risk, or lack of transportation). Beginning in July, we also asked participants if they completed COVID-19 serologic assessment and its outcomes. Also beginning in July, depression and anxiety were also assessed using short-forms developed from the Patient-Reported Outcomes Measurement Information System (PROMIS), which have been validated for populations living with chronic conditions;^12–15^ raw responses to questionnaires were converted to T-scores, which have a population mean of 50 and standard deviation of 10, with higher scores indicating greater depression or anxiety.

### Assessment of COVID-19 disease

In each survey, participants were asked several COVID-19-related questions including whether 1) a health provider ever suspected them of having COVID-19, 2) they had tested positive for COVID-19, or 3) had received a positive COVID-19 serology assessment. In addition, Johns Hopkins also maintains a COVID-19 registry in which all tests, results and COVID-19 outcomes performed within the state of Maryland or District of Columbia (via the Chesapeake Regional Information System for our Patients [CRISP], a regional health information exchange resource for Maryland/District of Columbia) are automatically uploaded into a database designed for research; we linked participants in COVID-RIMS with this registry to also allow for maximum case capture. Participants with self-reported positive COVID-19 tests by nasal swab RT-PCR or serum antibody testing, self-reported healthcare provider suspected COVID-19 (but were never tested) or had tested positive in CRISP were included as cases. We performed sensitivity analyses excluding individuals with suspected COVID-19 from the case definition.

### Statistical analysis

Initial analyses compared demographic characteristics of invited participants versus those who agreed to participate. Baseline demographics, disease characteristics, and key comorbidities were reported with descriptive statistics (e.g., mean and standard deviation [SD] for continuous variables and frequency and percentage for categorical variables). Initial analyses evaluated non-medication associated risk factors for COVID-19 and considered: age, sex, race, SES, working onsite, in-person socialization, smoking status, number of autoimmune or inflammatory conditions, number of immune-modulating medications exposures in the previous year and number of comorbidities using logistic regression models (as participants did not report exact timing of infection and we included participants with positive COVID-19 antibody testing in which exact timing of infection may be less clear). For in-person socialization, we considered both at baseline (when COVID-19 restrictions and business closures in Maryland and most of the United States were at their peak) and during follow-up, when restrictions had been somewhat relaxed. Comorbidities considered included hypertension, diabetes, cardiovascular disease (CVD; coronary heart disease, stroke, or heart failure), lung disease (chronic obstructive pulmonary disease [COPD], interstitial lung disease, or pulmonary hypertension), chronic kidney disease, asthma, and obesity (BMI ≥ 30). We also considered comorbidity burden as the sum of individual comorbidities affecting an individual. Primary analyses then assessed the association between exposure to different classes of immunomodulatory agents and risk of COVID-19 adjusted for age, sex, race, SES, working onsite, in person socialization, smoking status, number of autoimmune or inflammatory conditions and comorbidity also using logistic regression models. We categorized immunomodulatory/suppressive medications based on biologic class/mechanism or relative potency and considered the following categories: tumor necrosis factor (TNF)-inhibitors (e.g., adalimumab, infliximab, etanercept), B-cell depleting biologic agents (e.g., rituximab, ocrelizumab), other biologic therapies (e.g., natalizumab, abatacept, tocilizumab), conventional disease modifying drugs (DMDs; e.g., leflunomide, methotrexate, sulfasalazine, interferon-beta), hydroxychloroquine, strong immunosuppressants (e.g., cyclophosphamide), glucocorticoids, and intravenous immune globulin (IVIG) or plasmapheresis. We also assessed risk of COVID-19 associated with use of any biologic or non-biologic (e.g., collapsing the strong immunosuppressant and conventional DMDs categories). A full list of individual medications considered, and their associated category, are included in **eTable 7**. We also assessed risk of COVID-19 associated with medication classes stratified by disorders and individual medication exposures (as ever exposed, exposure within the past 1 year, exposure over 1 year ago) for medication classes or medications in which at least 10 COVID-19 cases were recorded. We also assessed predictors of interruptions to care (any interruption over follow-up, interruption to infusion, mental health or rehabilitative services, since these services are common in this population) using univariate and similarly adjusted multivariable-adjusted models performed with logistic regression methods. Lastly, we evaluated how depression and anxiety symptoms changed over the course of follow-up and evaluated predictors of higher overall symptom burden using univariate and multivariable-adjusted mixed effects models. Statistical calculations were performed with R software, version 3.6.2.^16^

## RESULTS

We initially invited 22,516 eligible patients, of whom 4666 (20.9%) agreed to participate and have completed at least one follow-up survey as of December 2020. Multiple sclerosis (MS), Sjogren’s syndrome, and rheumatoid arthritis were the most common autoimmune or inflammatory conditions affecting participants with 878, 741, and 545 individuals, respectively **(eTable 1)**. COVID-RIMS participants were more likely to be female (76.7% vs. 72.3%), white (83.1% vs. 63.2%) and have higher SES (mean ADI [SD] 23.7 [20.4] vs. 31.2 [24.4]) relative to those who did not respond to our survey invitation **(eTable 2)**. COVID-RIMS participants completed weekly (and later monthly) surveys for the duration of follow-up for a total of 10 surveys (baseline + 9 follow-up surveys); the median number of follow-up surveys completed was 8 (IQR: 5-9).

**Table 2.** Association between participant characteristics and COVID-19 risk in COVID-RIMS participants.

|  | Univariate |  | Multivariable* |  |
| --- | --- | --- | --- | --- |
|  | OR (95% CI) | P value | OR (95% CI) | P value |
| Age (per 10y) | 0.85 (0.78, 0.93) | 0.0004 | 0.84 (0.76, 0.93) | 0.0009 |
| Male sex | 0.76 (0.55, 1.04) | 0.08 | 0.82 (0.59, 1.14) | 0.55 |
| Race |  |  |  |  |
| White | 1.00 [ref] |  | 1.00 [ref] |  |
| Asian | 0.42 (0.13, 1.33) | 0.14 | 0.38 (0.12, 1.23) | 0.12 |
| Black/African American | 1.40 (0.96, 2.04) | 0.08 | 1.28 (0.86, 1.89) | 0.19 |
| Other | 1.06 (0.55, 2.03) | 0.87 | 1.01 (0.52, 1.97) | 0.86 |
| Unknown | 1.22 (0.37, 3.96) | 0.74 | 1.33 (0.41, 4.39) | 0.64 |
| Low SES (<25th percentile of ADI) | 1.48 (1.03, 2.13) | 0.04 | 1.38 (0.94, 2.01) | 0.10 |
| Number of autoimmune/inflammatory conditions | 1.16 (0.98, 1.37) | 0.09 | 1.17 (0.98, 1.39) | 0.08 |
| Ever exposed to an immune modulating agent | 1.61 (1.00, 2.59) | 0.05 | 1.44 (0.84, 2.46) | 0.18 |
| Number of autoimmune immune modulating agents exposed to in past year |  |  |  |  |
| 0 | 1.00 [ref] |  | 1.00 [ref] |  |
| 1 | 1.08 (0.79, 1.47) | 0.65 | 0.91 (0.64, 1.29) | 0.59 |
| 2 | 1.32 (0.92, 1.90) | 0.13 | 1.09 (0.74, 1.61) | 0.66 |
| 3+ | 1.54 (1.05, 2.24) | 0.03 | 1.20 (0.79, 1.81) | 0.40 |
| Obesity | 0.88 (0.67, 1.17) | 0.38 | 0.81 (0.61, 1.08) | 0.15 |
| Number of comorbidities** | 1.10 (0.99, 1.22) | 0.08 | 1.19 (1.06, 1.33) | 0.002 |
| Working onsite | 1.87 (1.44, 2.42) | <0.0001 | 1.69 (1.28, 2.22) | <0.0001 |
| Current smoker | 0.83 (0.40, 1.71) | 0.61 | 0.76 (0.37, 1.59) | 0.47 |
| Socializing in person at baseline | 1.13 (0.83, 1.54) | 0.445 | 1.01 (0.73, 1.38) | 0.97 |
| Socializing in person at any point in follow-up | 1.47 (1.13, 1.92) | 0.004 | 1.45 (1.10, 1.91) | 0.94 |

\*mutually adjusts for all variables included in the table
\*\*includes diabetes, CVD, lung disease (COPD, interstitial lung disease, pulmonary hypertension), stroke, asthma, CKD, hypertension, cancer

**Table 1** summarizes characteristics of responders overall and by COVID-19 status; 2187 (46.7%) report having been tested at least once for COVID-19 and 265 (5.7%) individuals reported positive COVID-19 results over the course of follow-up. For all responders, both those who developed COVID-19 and those who did not, 4161 (89.1%) reported ever being treated with an immune-modulating/suppressing medication, 167 (3.6%) were smokers, 1344 (28.8%) were obese, and 2736 (58.6%) had at least one medical comorbidity potentially associated with more severe COVID-19 disease or COVID-19 related hospitalization (e.g. hypertension, diabetes, CVD, lung diseases, chronic kidney disease [CKD], stroke, cancer); of the medical comorbidities considered, hypertension (30.8%) and CVD (18.8%) were most common.

Some patient characteristics were associated with differences in social distancing behaviors. For example, younger individuals, men, non-white individuals, those with lower SES, those with a higher comorbidity burden and those with exposure to a greater number of immune modulating/suppressing agents were less likely to socialize in person over follow-up **(eTable 3)**. For example, individuals exposed to 3+ immune modulating/suppressing agents in the past year were 20% less likely to socialize in person relative to those who did not report being on immunosuppression or immunomodulation medications in the past year (OR: 0.80; 95% CI: 0.65, 0.97). In contrast, individuals working on site were over two-fold more likely to report socializing in person (OR: 2.28; 95% CI: 1.96, 2.65).

**Table 3.** Association between patient characteristics and disruption to routine healthcare or related services

|  | Any disruption in services* (975 of 2156) |  |  |  |
| --- | --- | --- | --- | --- |
|  | Univariate |  | Multivariable** |  |
|  | OR (95% CI) | P value | OR (95% CI) | P value |
| Age, per 10 years | 1.01 (0.95, 1.08) | 0.659 | 1.03 (0.96, 1.11) | 0.355 |
| Male sex | 0.73 (0.59, 0.89) | 0.003 | 0.79 (0.64, 0.99) | 0.039 |
| Low SES | 0.99 (0.74, 1.33) | 0.970 | 0.94 (0.69, 1.29) | 0.718 |
| Race |  |  |  |  |
| White | 1.00 [ref] | - | 1.00 [ref] | - |
| Asian | 1.12 (0.60, 2.10) | 0.725 | 1.14 (0.59, 2.20) | 0.706 |
| Black/African American | 1.12 (0.83, 1.52) | 0.449 | 1.03 (0.75, 1.43) | 0.843 |
| Other | 1.33 (0.85, 2.10) | 0.209 | 1.23 (0.77, 1.97) | 0.388 |
| Unknown | 0.82 (0.34, 2.03) | 0.675 | 0.82 (0.32, 2.10) | 0.677 |
| Number of comorbidities*** | 1.10 (1.03, 1.19) | 0.008 | 1.07 (0.99, 1.16) | 0.097 |
| Obesity | 0.87 (0.72, 1.04) | 0.13 | 0.89 (0.73, 1.08) | 0.229 |
| Moderate to severe anxiety | 1.64 (1.32, 2.03) | <0.0001 | 1.53 (1.20, 1.94) | 0.0004 |
| History of depression | 1.06 (0.88, 1.27) | 0.556 | 0.92 (0.75, 1.13) | 0.443 |
| Number of autoimmune diagnoses | 1.12 (0.99, 1.27) | 0.073 | 1.06 (0.93, 1.21) | 0.374 |
| Ever treated with an immune-modulating medication | 1.03 (0.77, 1.37) | 0.853 | 0.99 (0.73, 1.35) | 0.95 |
| Working onsite | 0.71 (0.57, 0.87) | 0.001 | 0.71 (0.57, 0.90) | 0.004 |
| In person socializing over follow-up | 0.88 (0.70, 1.10) | 0.263 | 0.95 (0.75, 1.20) | 0.667 |
| In person socializing at baseline | 0.80 (0.68, 0.96) | 0.014 | 0.86 (0.71, 1.04) | 0.114 |
| COVID-19 infection | 1.52 (1.06, 2.16) | 0.022 | 1.35 (0.93, 1.96) | 0.115 |
| Change in ability to pay for disorder associated costs | 2.26 (1.80, 2.84) | <0.0001 | 1.96 (1.54, 2.50) | <0.0001 |
| Change in employment status due to COVID-19 | 1.48 (1.15, 1.90) | 0.002 | 1.34 (1.02, 1.76) | 0.034 |

\*includes any disruption to infusions, mental health, or rehabilitative services.
\*\*mutually adjusts for all variables included in the table
\*\* includes diabetes, CVD, lung disease (COPD, interstitial lung disease, pulmonary hypertension), stroke, asthma, CKD, hypertension, cancer

### Risk factors for contracting COVID-19

Younger age, comorbidity burden, working onsite and in-person socialization were each associated with increased odds of COVID-19 in multivariable-adjusted models **(Table 2)**. Individual comorbidities including diabetes (OR: 1.72; 95% CI: 1.08, 2.73), CVD (OR: 1.68; 95% CI: 1.24, 2.28), CKD (OR: 1.76; 95% CI: 1.04, 2.97) were also associated with COVID-19 odds in multivariable-adjusted models **(eTable 4)**. Results were similar when stratified by disorder; no significant heterogeneity when pooling results across disorder for estimated ORs.

**Table 4.** Patient characteristics associated with overall anxiety and depressive symptom burden

| Characteristic | Anxiety |  | Depression |  |
| --- | --- | --- | --- | --- |
|  | Mean difference in symptoms* (95% CI) | P value | Mean difference in symptoms* (95% CI) | P value |
| Age, per 10 years | -1.37 (-1.64, -1.10) | <0.0001 | -0.68 (-0.92, -0.43) | <0.0001 |
| Male sex | -3.44 (-4.19, -2.68) | <0.0001 | -1.06 (-1.75, -0.38) | 0.002 |
| Race |  |  |  |  |
| white |  |  |  |  |
| Asian | -1.54 (-3.34, 0.26) | 0.093 | -0.52 (-2.15, 1.12) | 0.536 |
| Black/African American | -1.84 (-2.94, -0.73) | 0.001 | -1.11 (-2.11, -0.11) | 0.03 |
| Other | 0.17 (-1.49, 1.82) | 0.844 | 0.06 (-1.45, 1.57) | 0.936 |
| Unknown | 2.30 (-1.03, 5.62) | 0.176 | 1.45 (-1.57, 4.48) | 0.346 |
| Low SES | 0.81 (-0.31, 1.94) | 0.156 | 0.83 (-0.20, 1.85) | 0.114 |
| Number of comorbidities** | 0.29 (-0.04, 0.63) | 0.089 | 0.26 (-0.04, 0.57) | 0.094 |
| History of depression | 4.89 (4.09, 5.70) | <0.0001 | 5.97 (5.24, 6.70) | <0.0001 |
| Obesity | 0.14 (-0.55, 0.84) | 0.686 | -0.07 (-0.70, 0.56) | 0.83 |
| Change in immune-modulating therapy | 0.09 (-0.44, 0.62) | 0.735 | -0.02 (-0.49, 0.45) | 0.942 |
| In person socialization | -0.42 (-0.75, -0.09) | 0.014 | -0.42 (-0.71, -0.13) | 0.004 |
| Working onsite | -0.20 (-0.68, 0.28) | 0.413 | -0.28 (-0.71, 0.14) | 0.187 |
| Change in ability to pay for disorder-associated costs | 2.17 (1.33, 3.01) | <0.0001 | 2.45 (1.70, 3.20) | <0.0001 |
| Change in employment status due to COVID-19 | 0.73 (0.10, 1.35) | 0.023 | 0.80 (0.25, 1.34) | 0.004 |
| COVID-19 infection | -0.38 (-1.68, 0.91) | 0.562 | -0.84 (-2.02, 0.34) | 0.164 |

\*mean difference in T-scores for anxiety and depression are estimated from a mixed effect model allowing for multiple assessments per person and also adjusting for follow-up time (categorically as month of follow-up). All estimates are adjusted simultaneously for all other variables included in the table.
\*\* includes diabetes, CVD, lung disease (COPD, interstitial lung disease, pulmonary hypertension), stroke, asthma, CKD, hypertension, cancer

When considering immune medications, glucocorticoid use in the past year was associated with 43% increased odds of COVID-19 **(Figure 2;** odds ratio [OR]: 1.43; 95% CI: 1.08, 1.89) in multivariable-adjusted models. With respect to glucocorticoid dose (as the sum of prednisone, methylprednisolone, and dexamethasone exposure), individuals reporting daily prednisone equivalent doses of 0.5 to 7.5mg/day, 7.5 to 60 mg/day and >60 mg/day had respective ORs of COVID-19 of 1.57 (95% CI: 1.06, 2.34), 1.60 (95% CI: 1.05, 2.44), and 1.95 (95% CI: 0.87, 4.39) relative to those with no exposure. **(eTable 5)**. Results were relatively consistent in analyses stratified by disorder **(eTable 6)**. Beyond glucocorticoids, other medication classes did not appear to be associated with COVID-19 risk. In sensitivity analyses, when we reclassified sphingosine-1-phosphate inhibitors (S1P; ozanimod, siponimod, fingolimod) as strong (rather than conventional) immunosuppressive agents or reclassified mycophenolate mofetil, azathioprine, or mercaptopurine as conventional (rather than strong) DMD, it did not alter the findings (for S1P: OR: 1.10; 95% CI: 0.81, 1.50; for mycophenolate mofetil: OR: 1.14; 95% CI: 0.73, 1.77; for azathioprine: 1.19; 95% CI: 0.81, 1.73; for mercaptopurine: 1.13; 95% CI: 0.80, 1.60). Likewise, removing natalizumab or vedolizumab from biologic therapies (as these therapies may not affect peripheral immune responses) also did not alter findings (OR: 1.14; 95% CI: 0.73, 1.77). With the exception of prednisone use in the past year (OR: 1.67; 95% CI: 1.21, 2.30), we did not find strong evidence of higher odds of COVID-19 associated with other individual medications for which ≥10 COVID-19 cases occurred among users **(eTable 7)**.

**Figure 1.**
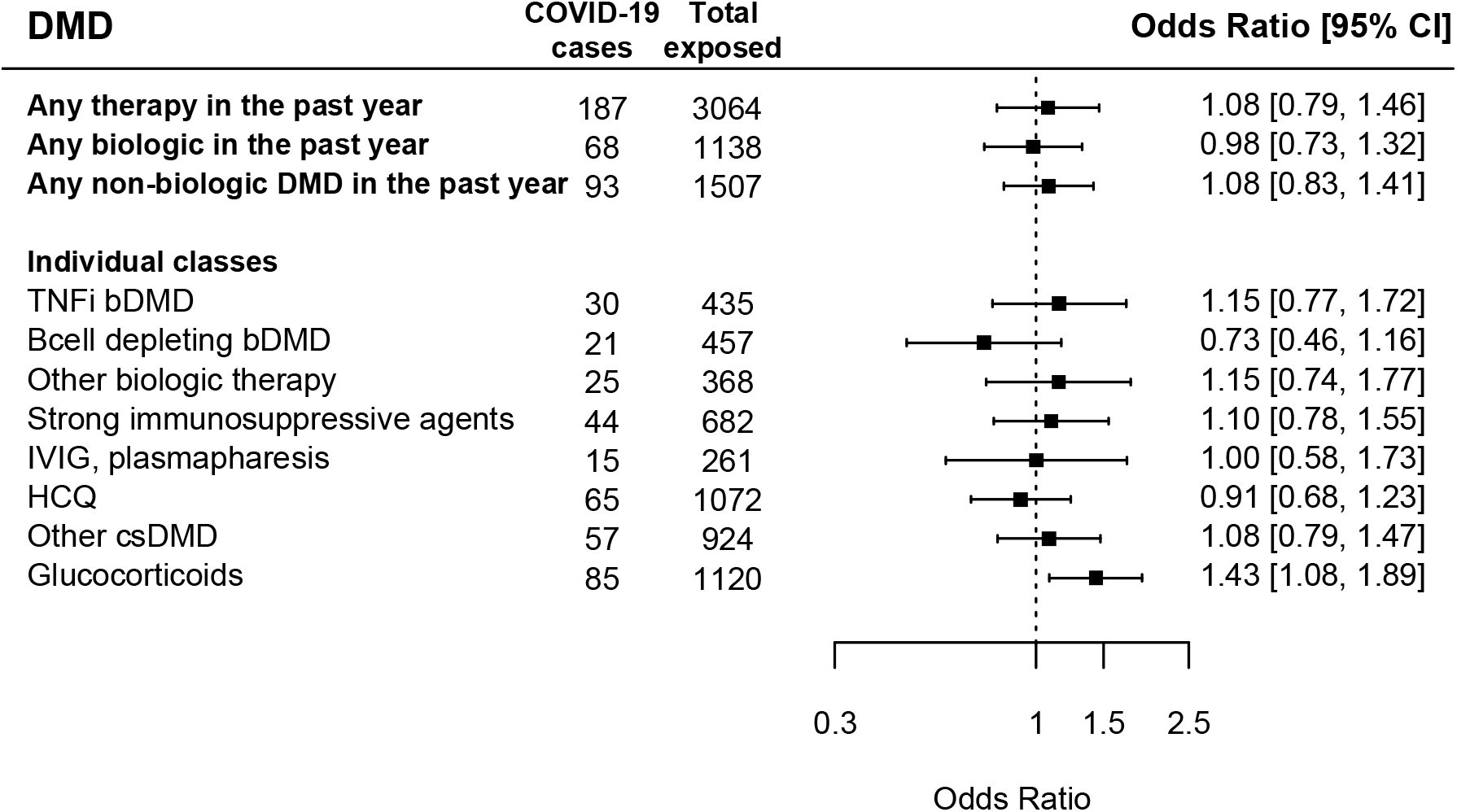
Association between immune-modulating or suppressive medications* and risk of COVID-19. DMD: disease modifying drug. Odds ratios are adjusted for age, sex, race, SES, working on site, in person socialization habits (at baseline and during follow-up), smoking status, number of comorbidities, number of autoimmune or inflammatory condition diagnoses, and current smoking status *For individuals medications included in each medication class, please refer to **eTable 7**.

**Figure 2.**
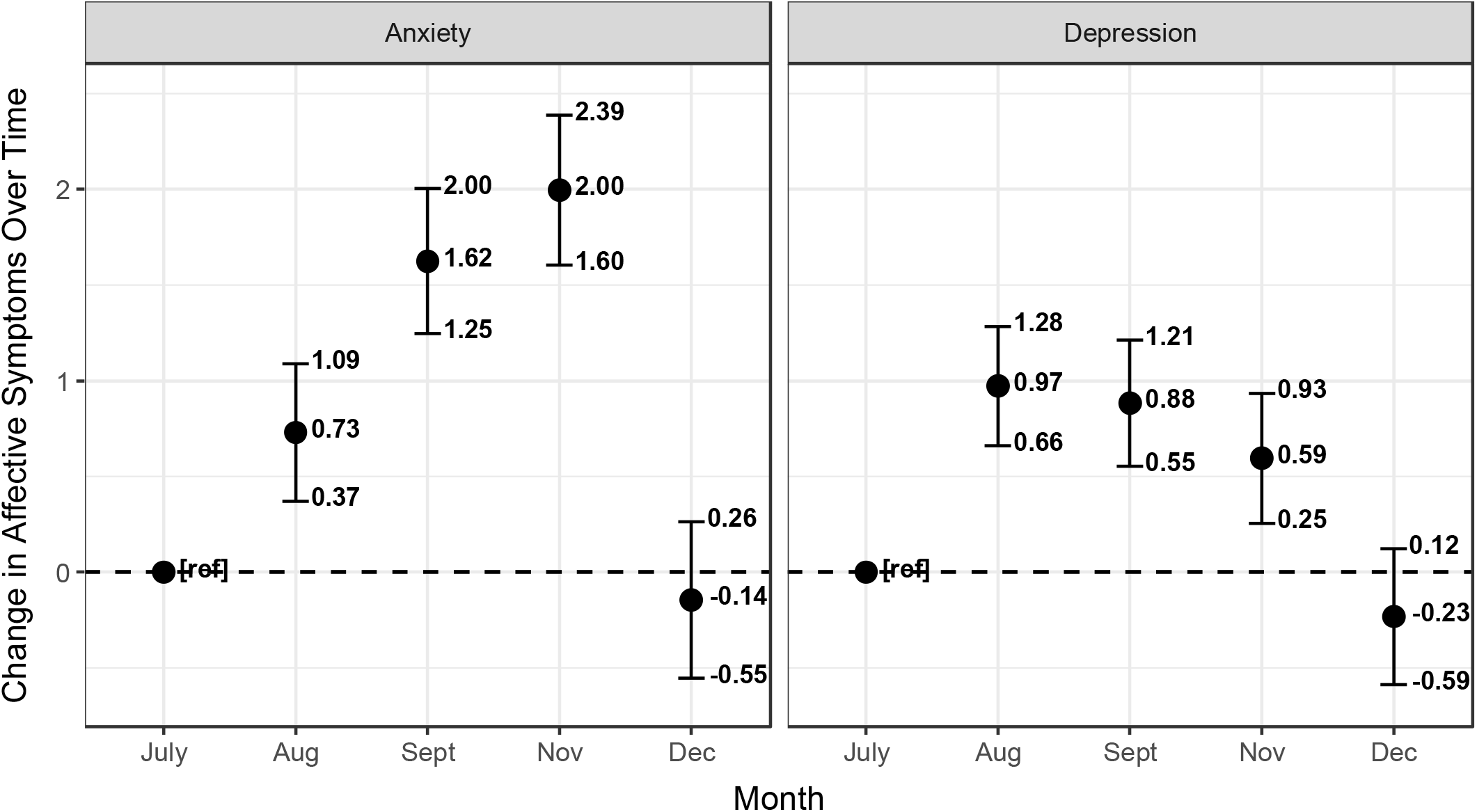
Change in anxiety (left) and depression (right) occurring over the course of study follow-up. Mean differences are adjusted for age, sex, race, SES, working on site, in person socialization, number of comorbidities, number of autoimmune or inflammatory condition diagnoses, COVID-19 infection, change in employment due to COVID-19, and changes in ability to pay for disorder-associated costs.

Assessment of risk factors for COVID-19 were consistent when excluding individuals who were suspected of having COVID-19 but were never tested (50 individuals excluded, leaving 215 COVID-19+ cases eligible for these analyses).

### Interruptions to care or services

Of the 2156 individuals who reported receiving infusions, rehabilitative or mental health services prior to the pandemic, 942 (45.2%) experienced an interruption to any services during the pandemic; 341 of 1158 (29.4%) delayed infusions, 623 of 1081 (57.6%) had an interruption in rehabilitative service, and 211 of 731 (28.8%) had interrupted mental healthcare **(Table 3** and **eTable 8)**. Those who experienced a change in their ability to pay for disorder-associated costs were nearly 2-fold more likely to have an interruption in services. Similarly, COVID-19-related changes to employment (e.g., furlough or termination) were also associated with an increased risk of service interruption (OR: 1.34; 95% CI: 1.02, 1.76). Those with moderate to severe anxiety (as defined as >1 SD above the mean T-scores from PROMIS measures) were 53% more likely to report an interruption in services (OR: 1.53; 95% CI: 1.20, 1.94). Men were less likely to experience an interruption in services (OR: 0.79; 95% CI: 0.64, 0.99). Findings were generally similar when considering specific reasons for disruption to services (e.g., infusions, rehabilitative services, or mental health).

### Overall burden of anxiety and depression and change over follow-up

On average, participants reported anxiety and depressive symptom T-scores of 51.8 [9.5] and 49.9 [8.5], respectively. Younger participants, men, and Black/African Americans tended to report a lower burden of anxiety and depression over the course of follow-up (e.g., lower T-scores), while previous self-reported previous physician-diagnosed depression was associated with substantially higher anxiety and depression symptom burden **(Table 4)**. Several pandemic-associated factors were also associated with higher symptom levels. For example, pandemic-associated changes in ability to pay for disorder-associated costs were associated with 2.17 (1.33, 3.01) points higher anxiety and 2.45 (1.70, 3.20) points higher depressive symptom T-scores. COVID-19 related changes to employment were also associated with small increases in anxiety and depressive symptoms, though it’s possible these changes may not be clinically meaningful (for anxiety: 0.73 points higher; 95% CI: 0.10, 1.35; for depression: 0.80; 95% CI: 0.25, 1.34).

Over the course of follow-up, the burden of depression and anxiety symptoms changed non-linearly. Peak anxiety levels occurred in November **(Figure 2;** 2.00 points higher; 95% CI: 1.60, 2.39). Depressive symptoms peaked slightly in August (0.97 points higher; 95% CI: 0.66, 1.28), though this difference may not be clinically meaningful. Changes in anxiety and depression occurring over follow-up were similar across different conditions and across different medication classes.

## DISCUSSION

In this study, we assessed risk factors for COVID-19 among a large cohort of patients with known autoimmune or inflammatory conditions in a well-phenotyped prospective registry at Johns Hopkins. Adherence to in person socialization recommendations were non-uniformly distributed across patients. Consistent with existing evidence, individuals working onsite or who are socializing in-person had a higher risk of contracting SARS-CoV-2 and developing COVID-19 disease. In addition, having diabetes, CKD, or CVD were associated with higher COVID-19 risk. With respect to immunomodulatory/suppressive medications, any glucocorticoid use in the past year was associated with an increased risk of COVID-19; other medications classes or individual medications themselves did not appear to be associated with COVID-19 risk. Our results were consistent in sensitivity analyses, where we varied both definitions for medication exposure as well as COVID-19 outcomes. Beyond COVID-19 disease, general interruptions to healthcare were common; individuals who experienced changes in their ability to pay for disorder-associated costs as well as those who experienced a COVID-19-related change to employment were most vulnerable to care disruptions. Overall, these findings suggest that as the COVID-19 pandemic continues to cause considerable morbidity and mortality, people with autoimmune or inflammatory disorders may be a particularly vulnerable population.

Results suggest that exposure to glucocorticoids may increase risk of contracting COVID-19. We did note a potential non-linear association between total glucocorticoid dose and COVID-19 risk. This observation is notable as glucocorticoids were among the most common medications used in this population, and many previous studies suggest a link between chronic exposure to glucocorticoids exposure and infection risk in people with autoimmune or inflammatory disorders.^17–20^ Also notable was our finding that other common medication classes were not strongly associated with COVID-19 infection. While it’s likely our sample size and number of identified COVID-19 cases precluded us from identifying the true COVID-19 risk associated with all of the DMDs considered, strong signals were not identified for more broad classifications of many of the common DMDs in this population. In the MS population, some prior studies had indicated an association between anti-CD20 therapy use and risk of COVID-19; these studies, while accounting for comorbidities, did not account for social distancing behaviors, which as we observed were non-uniformly distributed across patients.^1,2,7,21^ At the same time, the populations studied (in Europe) may have other characteristics that underlie the different outcomes. Most publications evaluating risk in other autoimmune disease evaluated risk of more severe disease rather than illness as a whole or used study designs that were less optimal (e.g., cross-sectional, case-control).^8,22,23^ With the success of recent COVID-19 vaccine trials, a critical next step will be to determine if and how common immunomodulating/suppressive medications or specific medication classes (e.g., B-cell depleting therapies) affect vaccination response, as has been shown for other vaccines.^24,25^

Notably, nearly half of participants who reported receiving infusion, rehabilitative, or mental health services reported a pandemic-related disruption to care. These results set the stage for future studies assessing the downstream consequences of these changes to autoimmune or inflammatory disease-specific outcomes, especially, as certain subgroups of patients (e.g., those with changes to household income) may be particularly vulnerable to these potential effects.

Finally, we also note non-linear changes in the burden of depression and anxiety. The sharp decrease in the trend of symptom burden between November and December could be related to announcements of the success of large COVID-19 vaccine effectiveness trials, which occurred in this period. It is also worth noting that the burden of mental health comorbidities is generally higher in many autoimmune and inflammatory disease populations relative to the general population;^26–30^ extended periods of social isolation may exaggerate symptoms of depression and anxiety in an already vulnerable population, an important observation for providers to keep in mind when caring for individuals with these conditions.

Our study has a number of important strengths. Our study is relatively distinct from prior research studies of individuals taking immune medications, which have largely focused on risk factors for poor COVID-19 outcomes. We included a large population of nearly 5000 individuals with autoimmune or inflammatory conditions who are already followed by specialists at a large health system, and, thus, may limit some biases related to right censoring that may be inherent to other studies including only hospitalized patients. Importantly, our study was also longitudinal and included information from patients collected at 10 different time points over the course of follow-up. We also included assessments of social distancing behavior to incorporate into analyses to ensure that any observed differences in medication-associated risks were not driven by differences of behavior. We also performed several sets of sensitivity analyses in which we assessed how varying assumptions of our analyses could impact the findings and our conclusions. Lastly, surveys included information related to disruptions to care, which may be particularly notable as these concerns affected a substantial proportion of participating patients. To date, most reports have focused on recommendations from providers, whereas our study collected such information on the observed burden in patients, which we found to be non-uniform across different subgroups of patients.

A number of important limitations of our study are worth noting. First, we lacked detailed information on exact timing of COVID-19 infection, so could not incorporate this information into the analyses or assess potential time-varying risk factors or confounding. We also could miss potential COVID-19 cases if participants were loss to follow-up or were not tested for COVID-19 in the Maryland/District of Columbia area, although, follow-up was relatively complete with participants completing a median 8 of 9 follow-up surveys. Further, we used self-reported medications, comorbidities, COVID-19 testing and result status. We also did not collect detailed information on medication dosages (beyond glucocorticoids), which could possibly lead to misclassification. We also identified eligible participants using diagnostic codes, when it is possible that this information may inaccurately identify patients. Nonetheless, we required ≥2 codes for a specific disease/condition from providers in specific specialty departments to reduce potential misclassification. Results are also derived from individuals who responded to our initial survey invitation and may not apply to the larger group of patients; responders were more likely to be white and have higher SES, so it’s possible our results have underestimated the impact of the pandemic in vulnerable groups. Lastly, as for any observational study, the potential for unmeasured confounding cannot be eliminated.

## Conclusion

Our findings are in line with existing research studies suggesting that exposure risks are strong risk-factors for contracting COVID-19. Other risk factors include a high comorbidity burden or a previous exposure to glucocorticoids. Disruption to healthcare and important related services were common, and non-universally distributed across patients. Those with pandemic-related changes to income (largely those with lower SES) may be a particular vulnerable subgroup, but providers should be mindful of potential delays of infusion therapies and disruption to care in general caused by COVID-19.

## Supporting information

eTable

## Data Availability

Anonymized data used for this study may be available from the corresponding author on reasonable request, with the proper data sharing agreements in place.

## Acknowledgements

We would like to acknowledge the following COVID-RIMS investigators for their contributions to the study: Brendan Antichos, Peter Calabresi, Laura Cappelli, Reezwana Choudhury, Jonathan Chrispin, Andrea Corse, Dana DiRenzo, Mark Donowitz, Matthew Elrick, Lisa Fox, Thomas Grader Beck, Regina Greco, Lindsey Hayes, Laura Hummers, Mohammed Khoshnoodi, Michael Kornberg, Nicholas Maragakis, Justin McArthur, Brett McCray, Joanna Melia, Zsuzsanna McMahan, Joanna Melia, John Miller, Brett Morrison, Lyle Ostrow, Alyssa Parian, Florin Selaru, Philip Seo, George Stojan, Harikrishna Tandri, Fredrick Wigley, and Huimin Yu.

